# Exercise-linked serum proteomics reveals a modifiable pre-cancer continuum

**DOI:** 10.64898/2026.07.28.26359103

**Authors:** Parthiban Periasamy, Jorming Goh, Siok Ghee, Patrick Sitjar, Thamil Selvan Vaiyapuri, Wu Yang, Sandra Lim, Zhang Zewen, Liu Wenrui, Denise Goh, Harsha Gowda, Yap Yoon Sim, Daniel Tan, Alan A Cohen, Roger Ho, Darren Lim, Fabian Lim, Tamás Fülöp, Elaine Lim, Jia Gengjie, Joe Yeong

## Abstract

Exercise reduces cancer incidence, yet the circulating molecular intermediates linking physical activity to pre-cancer biology remain unknown. Within the Singapore Longitudinal Ageing Studies (n = 6,050; ClinicalTrials.gov NCT03405675), we performed pre-cancer screening based on prospective cancer-free status at baseline serum collection followed by confirmed cancer diagnosis during longitudinal follow-up, with pre-diagnostic samples collected a median 7.76 years before cancer-specific death. Using serum proteomics (>2,400 proteins) across 674 samples — healthy controls (n = 89), pre-cancer individuals (n = 148) and cancer individuals (n = 57) — alongside longitudinal serum samples from two exercise paradigms: long-term unstructured vigorous activity (n = 134) and a short-term supervised structured intervention (n = 56), we identified 52 exercise-responsive hit proteins (HITs) that distinguish healthy from pre-cancer states, map a graded healthy-to-pre-cancer proteomic continuum and shift longitudinally toward healthier profiles following both exercise exposures. Both exercise signatures robustly discriminate pre-cancer from healthy participants, with performance that is predominantly protein-driven and minimally augmented by clinical covariates. A shared three-protein overlap signature retains comparable discrimination, with directional concordance independently confirmed in ∼9,800 UK Biobank participants via Olink proteomics. Together, these findings establish a biologically coherent, replicable exercise-linked proteomic signature that maps the pre-cancer state across two independent paradigms and motivates prospective, adherence-monitored interventional studies to determine whether these exercise-induced proteomic shifts are causally protective against cancer.

## Main text

Cancer develops over years before clinical diagnosis^1^. This preclinical interval represents a window in which host-response biology is already detectable in blood, but conventional laboratory tests lack sensitivity for the subtle, coordinated molecular perturbations associated with early cancer risk^2^. Circulating proteomics captures integrated immune, vascular and metabolic biology that is mechanistically relevant to carcinogenesis and immune surveillance. Physical activity is robustly associated with lower cancer incidence and improved post-diagnosis outcomes^3–5^, yet the circulating molecular intermediates linking activity to the biology of the pre-diagnostic cancer interval have not been identified.

Here, we profiled longitudinal serum proteomes across two complementary exercise paradigms — long-term unstructured vigorous activity and a short-term supervised structured intervention — quantified by liquid chromatography–tandem mass spectrometry (LC–MS/MS; >2,400 proteins). Exercise-responsive HIT proteins were identified by paired t-tests within intervention and matched control arms under FDR-controlled thresholds. Four proteomic signatures were constructed (structured, unstructured, overlap and union) and projected onto a disease-state comparator dataset. Healthy controls (n = 89) and pre-cancer individuals (n = 148) were drawn from the Singapore Longitudinal Ageing Studies (SLAS; n = 6,050)^6,7^; pre-cancer individuals were identified by cross-referencing participant records with the Singapore death registry, selecting those who were cancer-free at baseline serum collection and subsequently received a confirmed cancer diagnosis during follow-up, with baseline serum samples collected a median 7.76 years before cancer-specific death. Cancer patients (n = 57) were independently recruited patients with established cancer diagnoses (Supplementary Fig. 1). Directional external concordance was assessed in ∼9,800 UK Biobank participants^8^ with Olink Explore plasma proteomics.

In the unstructured cohort, paired baseline and follow-up sera from long-term exercisers (n = 34 pairs) and non-exercisers revealed 18 exercise-responsive HIT proteins (Fig. 1a). Pathway enrichment of these HITs identified coordinated downregulation of haemostasis and coagulation-fibrinolysis modules, with concurrent enrichment of cholesterol efflux, HDL remodeling and lipoprotein metabolism pathways (Fig. 1b). Seven representative HITs — IC1 (complement C1 inhibitor), DMKN (dermokine), SAA4 (serum amyloid A4), FIBA (fibrinogen beta chain), APOC2 (apolipoprotein C-II), A1AT (alpha-1-antitrypsin) and ITIH4 (inter-alpha-trypsin inhibitor heavy chain H4) — were all decreased in long-term exercisers relative to non-exercisers (n = 34 pairs; paired t-test p < 0.05 each; Fig. 1c). Downregulation of IC1^9,10^ and A1AT^11^ is consistent with attenuated complement and contact activation; reduced FIBA^12^ and ITIH4^10^ reflects lower coagulation and acute-phase tone; and changes in APOC2^13^ and DMKN^14^ are consistent with lipoprotein remodeling and extracellular matrix homeostasis under sustained vigorous activity. Together, these data demonstrate a coordinated vascular-metabolic and haemostatic exercise-response program in long-term exercisers.

**Fig. 1.**
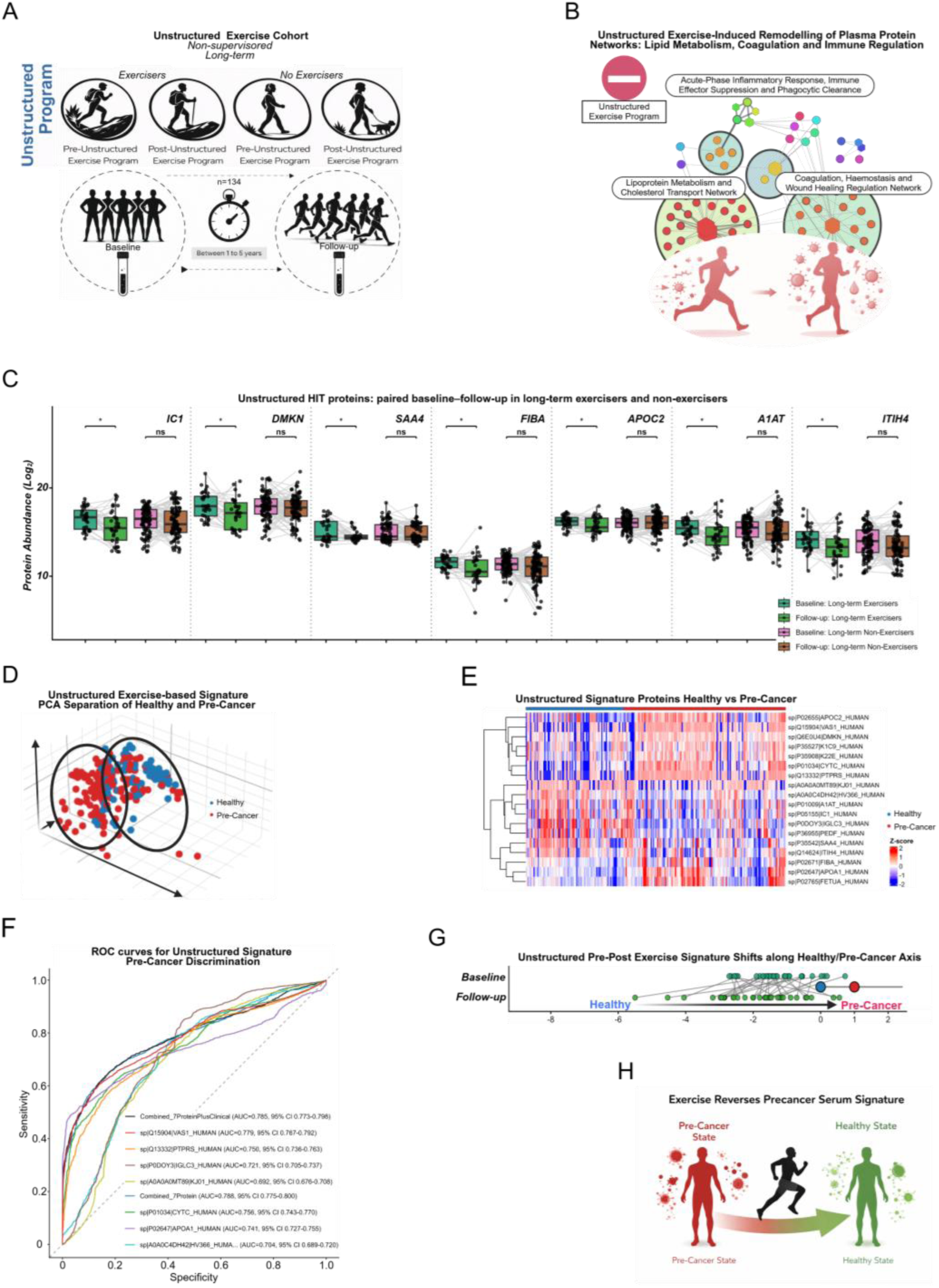
Long-term unstructured exercise remodels the plasma proteome toward a healthy state and discriminates pre-cancer from healthy individuals. a, Study design for the unstructured exercise cohort, with paired baseline and follow-up serum sampling in long-term exercisers and non-exercisers. b, Protein network diagram illustrating the functional modules remodeled by long-term unstructured exercise, including acute-phase inflammatory response, immune effector suppression and phagocytic clearance; coagulation, haemostasis and wound healing regulation; and lipoprotein metabolism and cholesterol transport. Node colour denotes functional cluster; edge weight reflects pathway connectivity. c, Paired boxplots of selected unstructured HIT proteins (IC1, DMKN, SAA4, FIBA, APOC2, A1AT and ITIH4) comparing baseline and follow-up abundance (log₂-scale) across long-term exercisers and non-exercisers. Brackets indicate paired t-test significance (*p < 0.05, ns = not significant). Each point represents one participant; lines connect paired timepoints. All seven proteins shown were significantly decreased in long-term exercisers relative to non-exercisers (paired t-test, p < 0.05 each). d, Three-dimensional principal component analysis (PCA) of the unstructured signature in healthy (blue) and pre-cancer (red) sera. e, Heatmap of row-wise z-scored unstructured-signature proteins in healthy and pre-cancer samples with hierarchical clustering. f, Receiver operating characteristic (ROC) curves for discrimination of pre-cancer versus healthy samples using the unstructured composite signature and individual proteins; area under the curve (AUC) values with 95% confidence intervals (CIs) are shown. g, Individual participant pre-to-post trajectories along the composite healthy–pre-cancer axis in the unstructured arm. h, Conceptual schematic of exercise-associated movement from pre-cancer-associated towards healthy-associated serum proteomic states.

The unstructured signature separated healthy from pre-cancer sera across three independent views: PCA (Fig. 1d), hierarchical heatmap architecture (Fig. 1e) and classifier performance (Fig. 1f). The combined unstructured model (18-protein unstructured composite), adjusted for age and sex, achieved a pre-cancer-versus-healthy AUC of 0.769 (95% CI, 0.755–0.782; Fig. 1f). At the individual level, longitudinal composite scores shifted toward the healthy pole at follow-up in exercisers (Fig. 1g), summarised schematically in Fig. 1h. Together, these data show that the unstructured exercise proteome is associated with movement of pre-cancer-associated circulating states toward healthy-like profiles.

In the structured intervention, 20 HIT proteins were identified from paired pre/post sera of supervised exercisers and waitlist controls (n = 40 pairs; Fig. 2a). Pathway enrichment showed coordinated upregulation of lipid-handling, HDL remodeling, adaptive immune and haemoglobin-related pathways (Fig. 2b). Seven representative HITs — HBA (α-haemoglobin), APOD (apolipoprotein D), RET4 (retinol-binding protein 4), VTDB (vitamin D-binding protein), HBB (β-haemoglobin), APOC2 (apolipoprotein C-II) and ATAG2 (ATG2A) — were all increased post-intervention in exercisers (n = 40 pairs; paired t-test p < 0.01 for HBA and APOD; Fig. 2c) but did not change significantly in waitlist controls. Upregulation of HBA^15^ and HBB^15^ is consistent with exercise-induced erythropoietic adaptation; APOD^16^, APOC2^13^ and RET4^17^ elevation reflects remodeling of lipoprotein particles and improved lipid-metabolic tone; VTDB^18^ upregulation may reflect enhanced vitamin D mobilisation; and ATAG2^19^ induction is consistent with activation of macroautophagy — a cellular quality-control program stimulated by exercise. Together, these data identify a coordinated metabolic, erythropoietic and proteostatic adaptation program specific to supervised structured training.

**Fig. 2.**
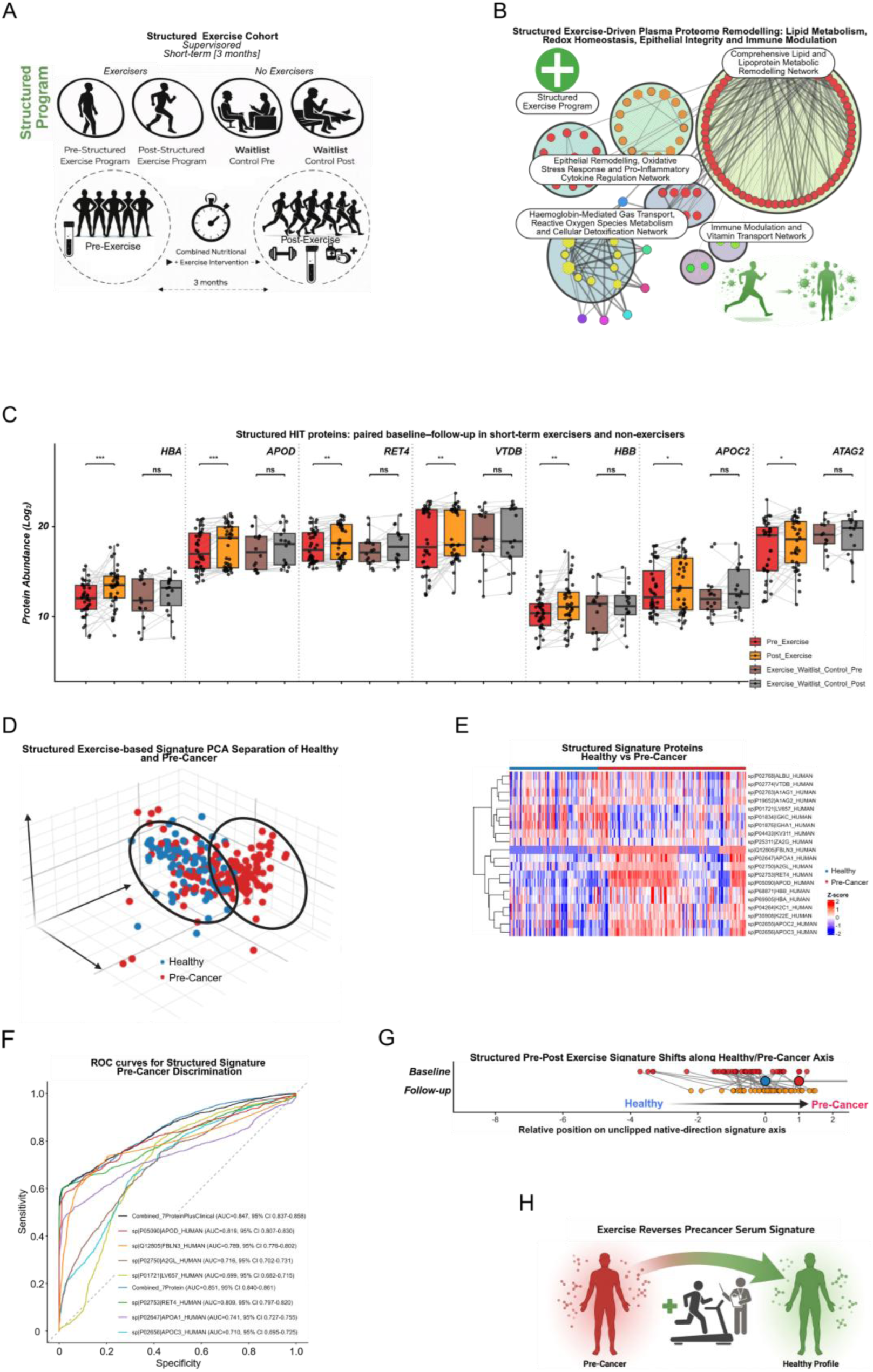
Short-term supervised structured exercise drives plasma proteome remodeling and shifts the pre-cancer protein signature toward a healthy profile. a, Study design for the supervised structured exercise intervention, with pre/post serum sampling in exercisers and comparator groups (waitlist control and waitlist-to-exercise). b, Protein network diagram of structured exercise-driven plasma proteome remodeling across four functional domains: comprehensive lipid and lipoprotein metabolic remodeling; haemoglobin-mediated gas transport, reactive oxygen species metabolism and cellular detoxification; epithelial remodeling, oxidative stress response and pro-inflammatory cytokine regulation; and immune modulation and vitamin transport. Node colour denotes functional cluster; edge weight reflects pathway connectivity. c, Paired boxplots of selected structured HIT proteins (HBA, APOD, RET4, VTDB, HBB, APOC2 and ATAG2) comparing pre- and post-intervention abundance (log₂-scale) in exercisers and waitlist controls. Brackets indicate paired t-test significance (*p < 0.05, **p < 0.01, ***p < 0.001, ns = not significant). Each point represents one participant. All seven proteins shown were significantly increased post-intervention in exercisers (paired t-test, p < 0.05 for leading analytes) but did not change significantly in waitlist controls, confirming exercise-specificity of the response. d, Three-dimensional PCA of the structured signature in healthy and pre-cancer sera. e, Heatmap of row-wise z-scored structured-signature proteins with hierarchical clustering. f, ROC curves for pre-cancer versus healthy discrimination using the structured composite signature and individual proteins; AUC values with 95% CIs are shown. g, Individual participant pre-to-post trajectories along the composite healthy–pre-cancer axis in the structured arm. h, Conceptual schematic of exercise-associated movement towards a healthy-associated serum proteomic state.

Routine clinical and anthropometric features failed to distinguish healthy from pre-cancer participants, showing broad overlap across PCA, integrated heatmap and differential feature analyses (Supplementary Fig. 2a–c). Across organ-specific pre-cancer cohorts, most routine laboratory analytes overlapped broadly with healthy reference values (Supplementary Fig. 3), although immune-inflammatory indices (neutrophil-to-lymphocyte, platelet-to-lymphocyte and systemic immune-inflammation index) showed comparatively stronger between-group structure. Lead time from pre-cancer blood draw to cancer-specific death was highly variable by tumour type (Supplementary Fig. 4), supporting interpretation of the cancer patients as contextual malignant-state biology rather than a uniform progression endpoint.

Disease-state mapping with the structured signature paralleled the unstructured arm. Healthy and pre-cancer sera separated in three-dimensional PCA (Fig. 2d) and showed coordinated abundance differences in hierarchical heatmap analysis (Fig. 2e). In ROC analysis, the structured combined model (20-protein structured composite) adjusted for age and sex achieved AUC = 0.823 (95% CI, 0.812– 0.835), with comparable performance for the protein-only model (AUC = 0.823; 95% CI, 0.811–0.834) and leading individual analytes (APOD, AUC = 0.819; RET4, AUC = 0.806; Fig. 2f). Many participants shifted toward healthier composite-score positions post-intervention (Fig. 2g; Fig. 2h schematic). Pooling organ-specific pre-cancer groups increased power to detect shared systemic features at the cost of tissue-specific resolution; these signatures therefore represent host-state signals rather than tissue-of-origin classifiers. Together, these data indicate a graded healthy-to-pre-cancer proteomic continuum that structured exercise can remodel.

Given convergence across exercise modalities, we evaluated whether minimal shared and broader composite signatures retained discrimination. The three-protein overlap panel was first evaluated as an overlap signature. Unsupervised PCA, hierarchical heatmap structure and ROC analysis each separated healthy from pre-cancer participants (Supplementary Fig. 5a–c), with the three-protein composite achieving AUC ≈ 0.821 (95% CI, 0.809–0.832) and only a modest increment from adding clinical covariates (AUC ≈ 0.826; 95% CI, 0.815–0.837; Supplementary Fig. 5c). The broader combined signature (35-protein union of all structured and unstructured HITs) recapitulated the same group structure across PCA, heatmap and composite-score views (Supplementary Fig. 5d–f) and yielded comparable ROC performance (Supplementary Fig. 5g). Projection of cancer patient samples (n = 57) extended the healthy-to-pre-cancer gradient toward overt malignancy. Cross-platform comparison with Olink Explore proteomics showed concordant overlap-signature structure in PCA and heatmap views (Supplementary Fig. 6a, b). To confirm that our mass spectrometry findings were not platform-specific, we compared them against an independent Olink proteomic platform. The same proteins that changed with exercise in our study also changed in the same direction in the Olink data, and this agreement was greater than what would be expected by random chance. This cross-platform consistency supports the biological authenticity of the exercise-responsive protein signatures identified in this study (Supplementary Fig. 6c–d). Together, these data show that exercise-linked proteomic differences are biologically reproducible across panel sizes and platforms.

We then examined the biological pathways altered in the pre-cancer state and asked whether exercise-responsive proteins from Figs. 1 and 2 map onto these same pathways — directly testing whether exercise biology and pre-cancer biology are connected at the molecular level. In pre-cancer, proteins involved in lipid metabolism, blood coagulation and cellular signaling were elevated relative to healthy controls (Supplementary Fig. 7a). Strikingly, these are the same pathways that both structured and unstructured exercise remodel in the opposite direction — APOD, APOC2 and RET4, which are upregulated by structured exercise (Fig. 2b– c), belong to pathways that are dysregulated in pre-cancer, and APOC2 and FIBA, modulated by unstructured exercise (Fig. 1b–c), operate within the same biological networks. This directional opposition — exercise pushing these pathways one way, pre-cancer pushing them the other — represents the central molecular observation of this study. Conversely, proteins involved in immune surveillance, complement defense, extracellular-matrix maintenance and waste clearance were reduced in pre-cancer (Supplementary Fig. 7b). Some of these same proteins — IC1^20^, A1AT^11^ and ITIH4^10^ — were also reduced by long-term unstructured exercise (Fig. 1c). This co-directional reduction does not imply equivalence: in pre-cancer, immune suppression co-occurs with pro-coagulant lipid excess as a combined pathological signature, whereas exercise simultaneously attenuates that lipid and coagulation excess, producing a fundamentally different net biological state. The immune protein changes under exercise reflect normal physiological adaptation, not disease-associated suppression. Together, this bifurcated pre-cancer landscape — elevated lipid and coagulation programs alongside reduced immune clearance — is summarised in Supplementary Fig. 7c, and the exercise signatures are directionally consistent with partially countering the pro-cancer proteomic shifts. These observations support a model in which regular physical activity may attenuate early cancer-associated molecular programs; however, since the exercise cohorts comprised healthy cancer-free individuals and pre-cancer participants were not directly subjected to exercise, these findings establish biological plausibility rather than proof of protection. Prospective studies testing exercise intervention directly in pre-cancer populations are required to determine whether these proteomic shifts translate into reduced cancer risk. Cross-dataset validation confirmed that the same pathways suppressed in pre-cancer — immune defense, cytoskeletal organisation and cellular stress responses — are also suppressed in independent lung cancer tissue (PXD066683; Supplementary Fig. 8a), lung cancer serum (PXD034221; Supplementary Fig. 8b) and breast cancer B-cells (PXD052185; Supplementary Fig. 8c), confirming that the biological programs identified here are relevant across cancer contexts and strengthening confidence that exercise-linked proteomic remodeling operates along pathways with genuine cancer-state relevance.

Collectively, these analyses establish a biologically coherent and replicable proteomic signature of physical activity that maps onto the pre-cancer state and is modified by exercise across two independent paradigms. The finding that proteomic remodeling was directionally convergent across non-equivalent exercise modalities — differing in duration (years versus months), supervision (self-directed versus coached) and modality (habitual aerobic activity versus structured multimodal training) — argues for a shared systemic response program rather than a paradigm-specific artefact. The pre-cancer blood samples were drawn a median 7.76 years before cancer-specific death, demonstrating that the proteomic signal is detectable within a clinically meaningful pre-diagnostic window and that exercise-linked remodeling operates along pathways active long before any clinical presentation. Together, these observations are consistent with a model in which exercise shifts the circulating proteome in a direction that partially counters early cancer-associated molecular changes — a biologically plausible mechanism through which physical activity may reduce cancer risk.

Several limitations warrant acknowledgement. Pre-cancer ascertainment was retrospective, residual confounding cannot be excluded, and organ-specific pre-cancer groups were pooled, limiting tissue-specific resolution. The exercise cohorts were not powered for cancer-outcome endpoints; interpretation therefore remains restricted to proteomic remodeling rather than causal or prognostic inference. Notably, keratin proteins — specifically K22E (cytokeratin type II cytoskeletal 2 epidermal), K1C9 (cytokeratin type I cytoskeletal 9) and K2C1 (cytokeratin type II cytoskeletal 1) identified as statistically significant HITs in the HIT discovery analysis. Their susceptibility to laboratory contamination during sample handling is well established^21^, and they were therefore excluded from primary signature visualisation and the representative protein panels in Figs. 1 and 2. However, K22E independently satisfied FDR-controlled paired statistical thresholds in both the structured and unstructured exercise cohorts — two separate studies with distinct designs — and was therefore retained in the three-protein overlap count. Whether this double-independent replication reflects genuine exercise-associated epithelial remodeling, consistent pre-analytical handling differences between baseline and follow-up visits, or a combination of both cannot be resolved without orthogonal targeted validation. Keratin-based serum markers have established clinical utility in oncology (for example, CYFRA 21-1 for lung cancer detection^22^), and the possibility that exercise influences circulating epithelial protein shedding represents a biologically plausible but unconfirmed hypothesis. Prospective interventional studies with standardised activity quantification, adherence monitoring, pre-analytical quality controls and longitudinal cancer endpoints are needed to determine whether these exercise-linked proteomic shifts — including the keratin-associated signals — are causally protective, clinically actionable, or both.

## Data Availability

All data produced in the present study are available upon reasonable request to the authors.

## Supplementary figures

**Supplementary Fig. 1.**
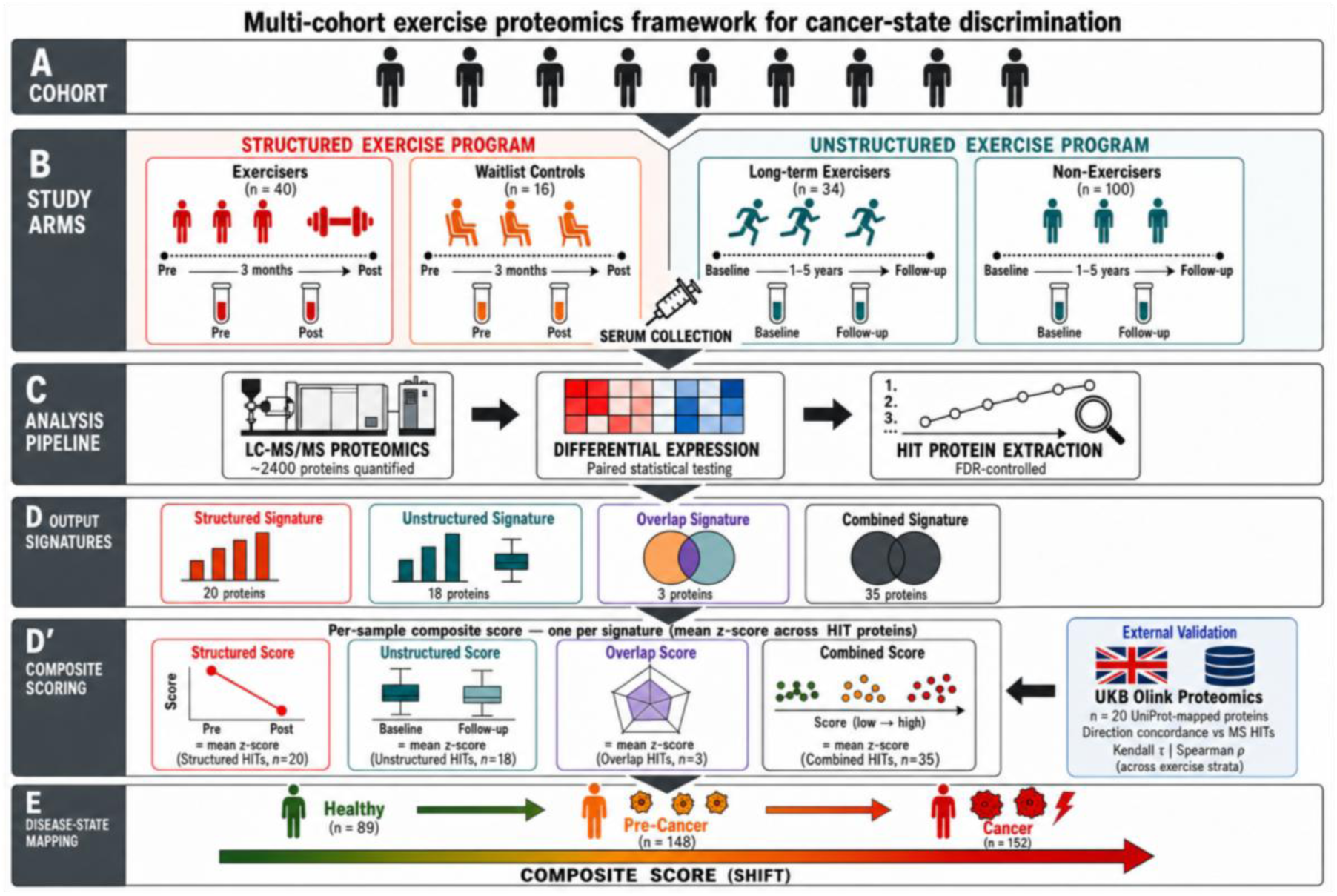
Study design and analytical framework. a, Multi-cohort overview: participants were drawn from a unified cohort and assigned to two independent exercise modalities for a parallel proteomic analysis. b, Study arms: the structured exercise arm comprised supervised exercisers and waitlist controls, with paired serum samples collected pre- and post-intervention (3 months); the unstructured exercise arm comprised long-term exercisers and non-exercisers, with paired serum samples collected at baseline and follow-up (1–5 years). c, Analysis pipeline: serum samples were profiled by liquid chromatography– tandem mass spectrometry (LC-MS/MS) proteomics quantifying approximately 2,400 proteins per sample; exercise-responsive proteins (HITs) were identified by paired t-tests within intervention and matched control arms; statistical significance was required in the intervention arm (paired t-test p < 0.05) with directional non-mirroring in the control arm, and hit sets were extracted under FDR-controlled thresholds. d, Four proteomic signatures were derived: a structured-exercise signature (20 proteins), an unstructured-exercise signature (18 proteins), an overlap signature (intersection of both; 3 proteins), and a combined signature (union of both; 35 proteins). d’, Composite scoring: a per-sample composite score was computed as the mean z-score across HIT proteins in each respective signature. Structured Score (n = 20 HITs, pre-versus-post slope), Unstructured Score (n = 18 HITs, baseline-versus-follow-up), Overlap Score (n = 3 HITs) and Combined Score (n = 35 HITs) were each computed independently. External validation was performed against UKB Olink proteomics (n = 20 UniProt-mapped proteins) using directional concordance (Kendall’s τ, Spearman’s ρ) across exercise strata. e, Each signature was mapped onto an independent disease-state cohort spanning healthy controls (n = 89), pre-cancer individuals (n = 148) and cancer patients (n = 57); a composite signature score axis quantifies the directional shift from a healthy-associated to a cancer-associated proteomic state.

**Supplementary Fig. 2.**
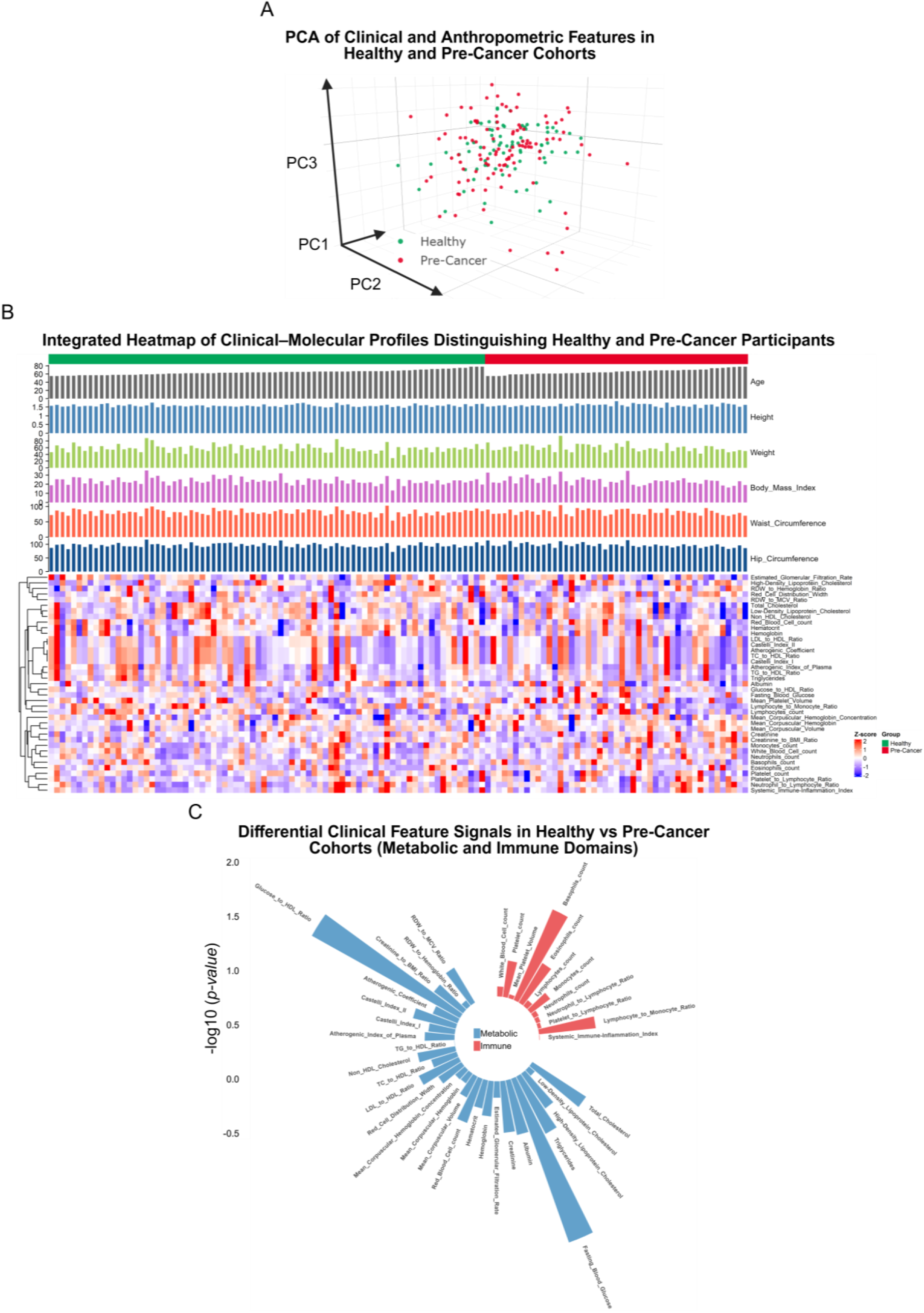
Clinical and anthropometric features alone are insufficient to distinguish healthy from pre-cancer individuals, underscoring the discriminatory value of the plasma proteome. a, Three-dimensional PCA of integrated clinical and molecular features showing healthy (green) and pre-cancer (red) samples. b, Integrated heatmap of clinical–molecular profiles across healthy and pre-cancer participants. Anthropometric annotation tracks (Age, Height, Weight, Body Mass Index, Waist Circumference, Hip Circumference) are shown above the molecular feature matrix; column annotation denotes disease group. Hierarchical clustering was applied to molecular features (rows). c, Radial summary plot of between-group differences shown as −log10(p-value) values across cardiometabolic and immune-inflammatory variables.

**Supplementary Fig. 3.**
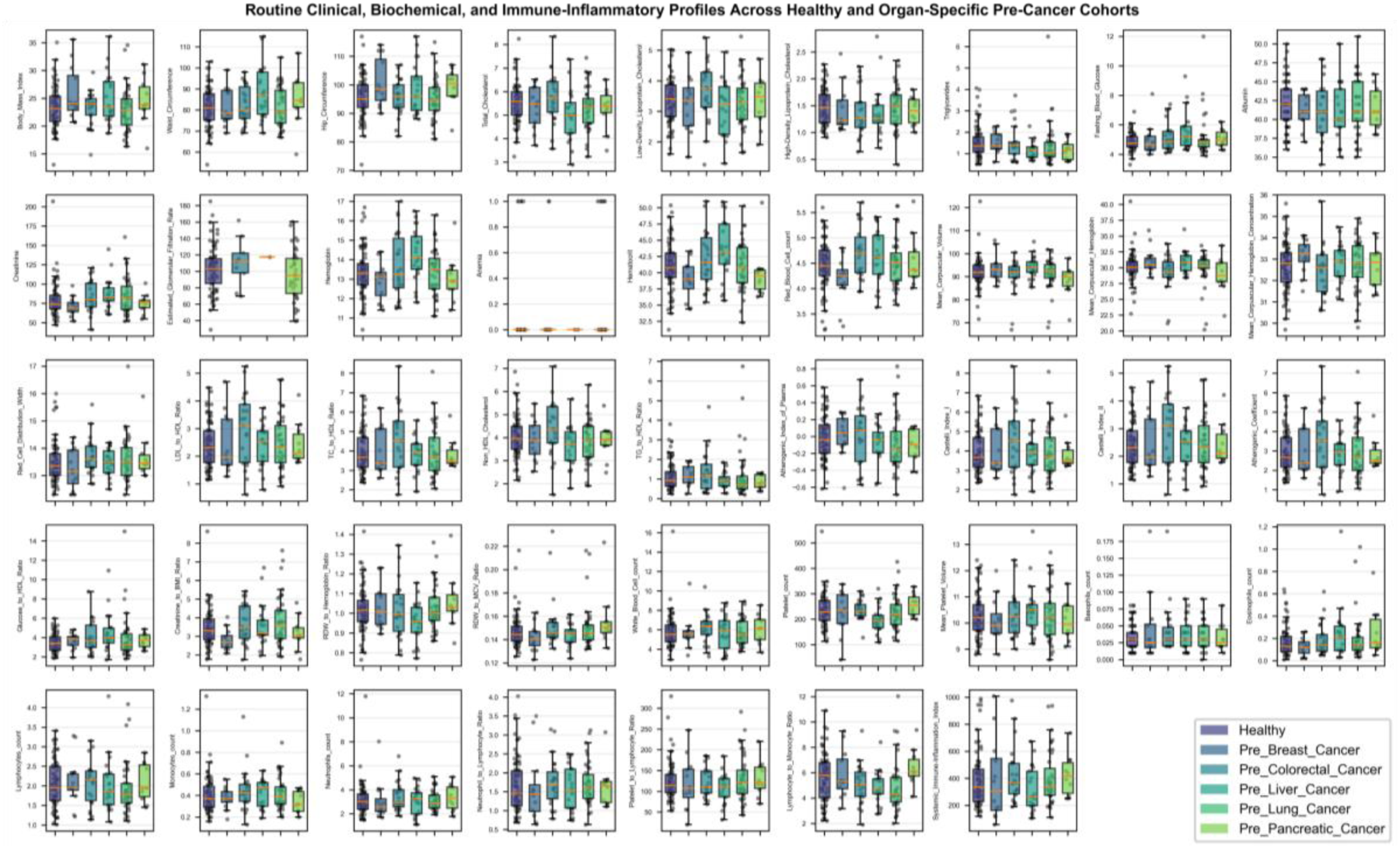
Routine clinical, biochemical and immune indices across healthy controls and organ-specific pre-cancer cohorts. Grid of box-and-whisker plots with overlaid individual data points comparing healthy participants with pre-breast, pre-colorectal, pre-liver, pre-lung and pre-pancreatic cancer groups across anthropometric, metabolic, renal/hepatic, hematologic and immune-inflammatory indices; significance annotations are shown where applicable.

**Supplementary Fig. 4.**
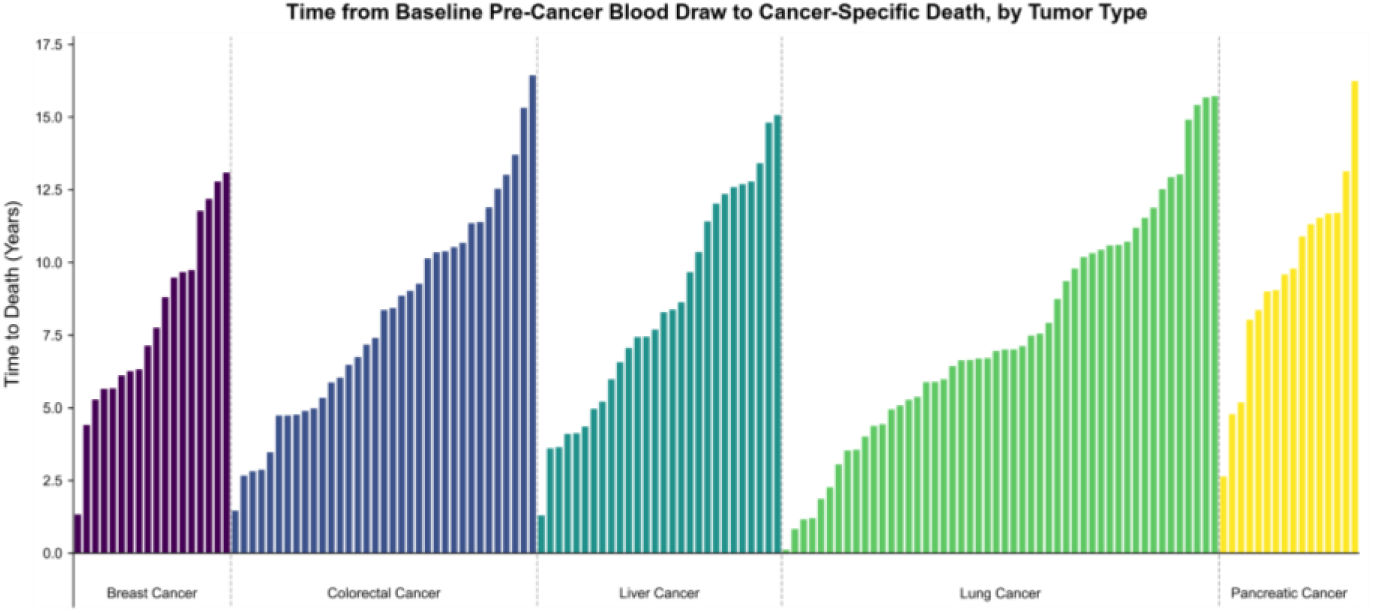
Time from baseline pre-cancer serum sampling to cancer-specific death, by tumor type. Waterfall plot showing participant-level time (years) from baseline serum collection to cancer-specific death for participants in the pre-cancer arm who were subsequently diagnosed with cancer and died of the disease; ordered within each tumor type (breast, colorectal, liver, lung and pancreatic).

**Supplementary Fig. 5.**
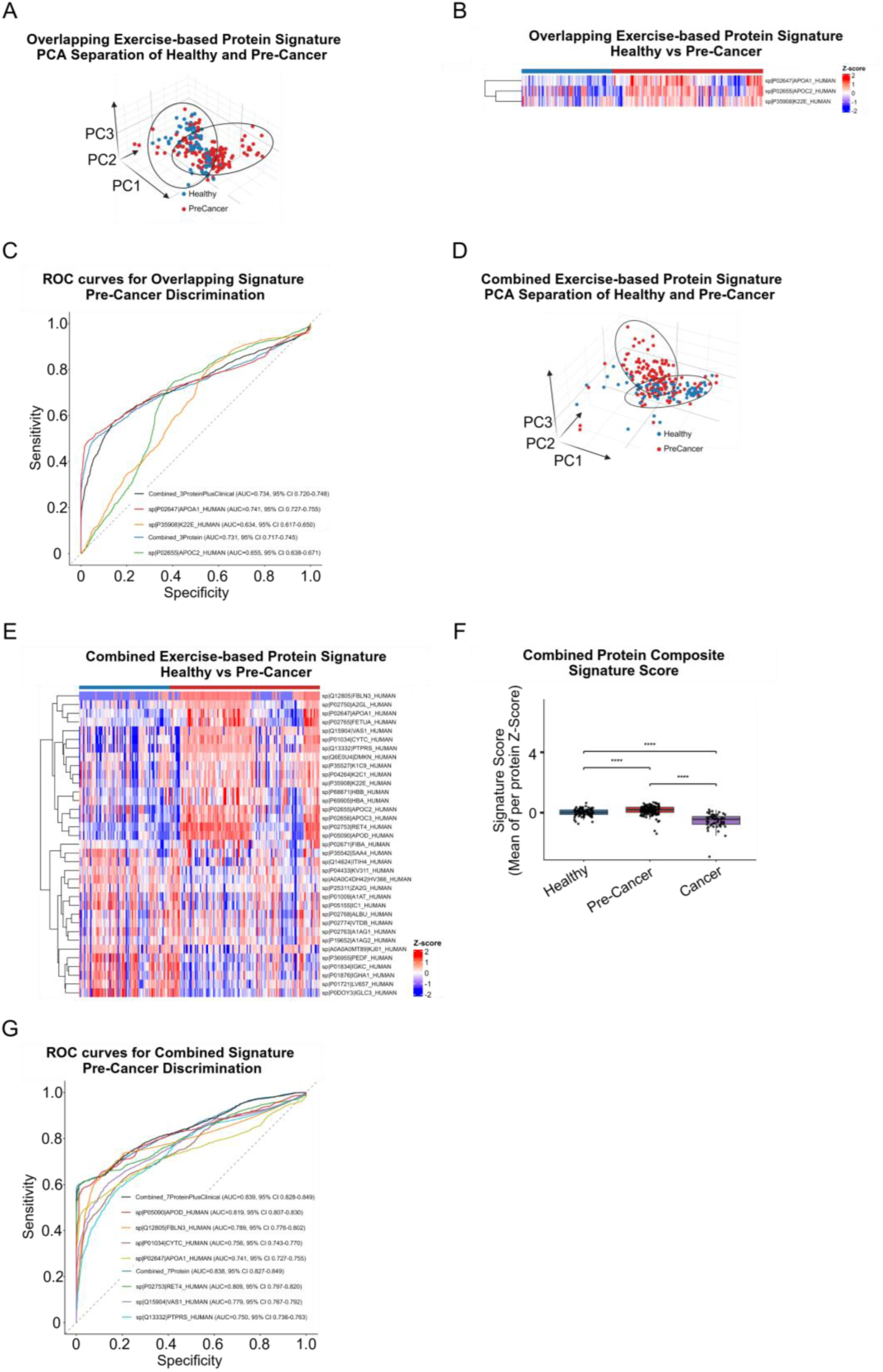
Overlapping and combined exercise-derived protein signatures retain discriminatory power for pre-cancer detection, with the combined composite score showing stepwise progression from healthy to pre-cancer to cancer. a–c, Overlap-signature analyses for the three overlap proteins (K22E, APOC2 and APOA1): PCA separation of healthy (blue) and pre-cancer (red) participants (a); heatmap of row-wise z-scored overlap signature proteins with hierarchical clustering (b); and ROC curves for pre-cancer-versus-healthy discrimination with AUC values and 95% confidence intervals (c). d–g, Combined-signature analyses (35-protein union signature): PCA separation of healthy and pre-cancer participants (d); heatmap of 35 combined exercise-based signature proteins (e); composite signature score boxplot across Healthy (n = 89), Pre-Cancer (n = 148) and Cancer (n = 57) groups with significance annotations (f); and ROC curves for pre-cancer-versus-healthy discrimination with AUC values and 95% confidence intervals (g).

**Supplementary Fig. 6.**
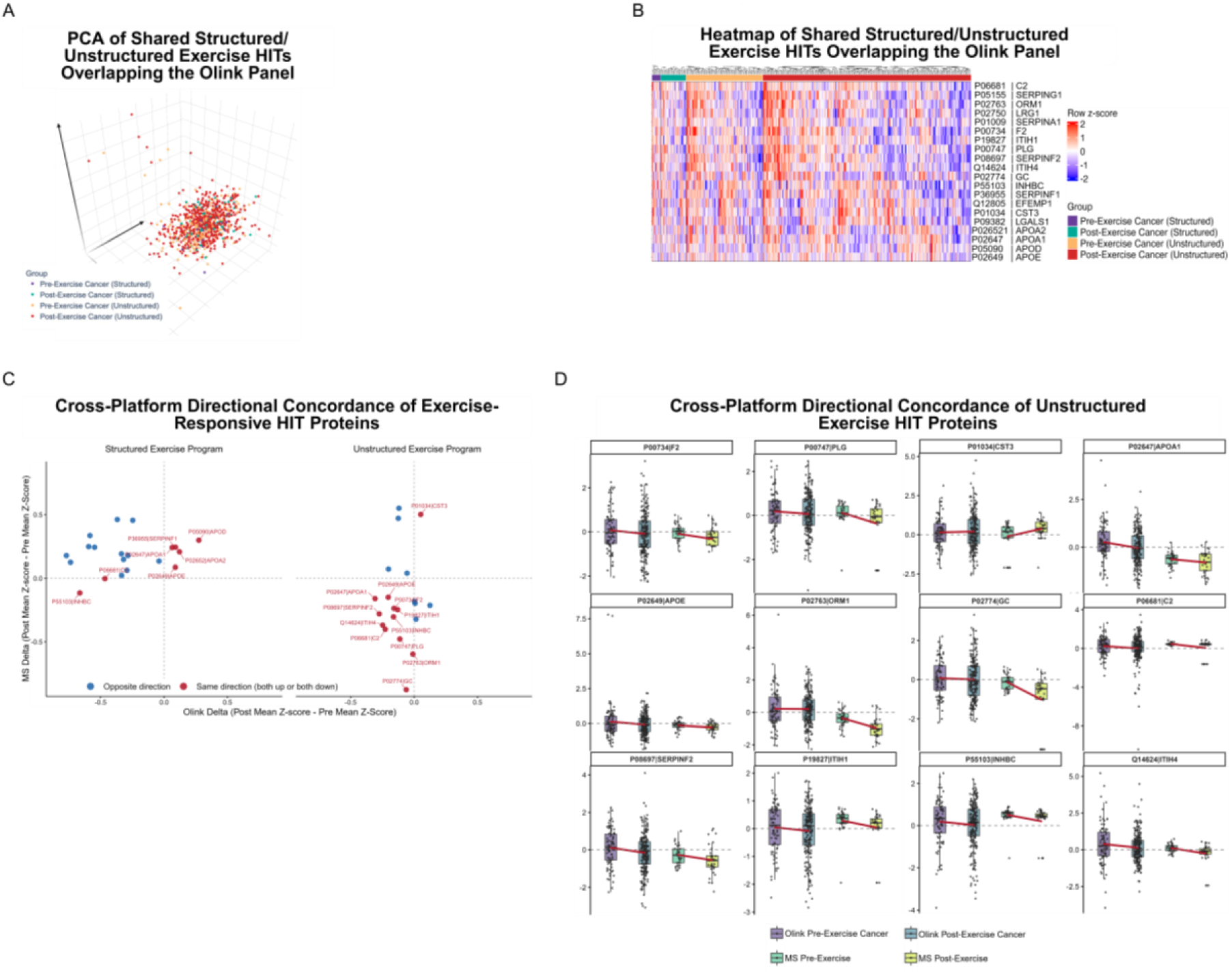
Olink overlap-signature structure and Olink–LC-MS/MS directional concordance across exercise modalities. a, PCA of Olink NPX data restricted to proteins overlapping structured and unstructured exercise HITs. b, Heatmap of the same Olink overlap panel (row-wise z-scored). c, Cross-platform directional concordance scatter comparing Olink mean Δ (post minus pre) and LC-MS/MS paired Δ for UniProt-matched analytes. d, Protein-level cross-platform pre/post boxplots in the unstructured arm illustrating direction-level concordance despite platform-specific scaling.

**Supplementary Fig. 7.**
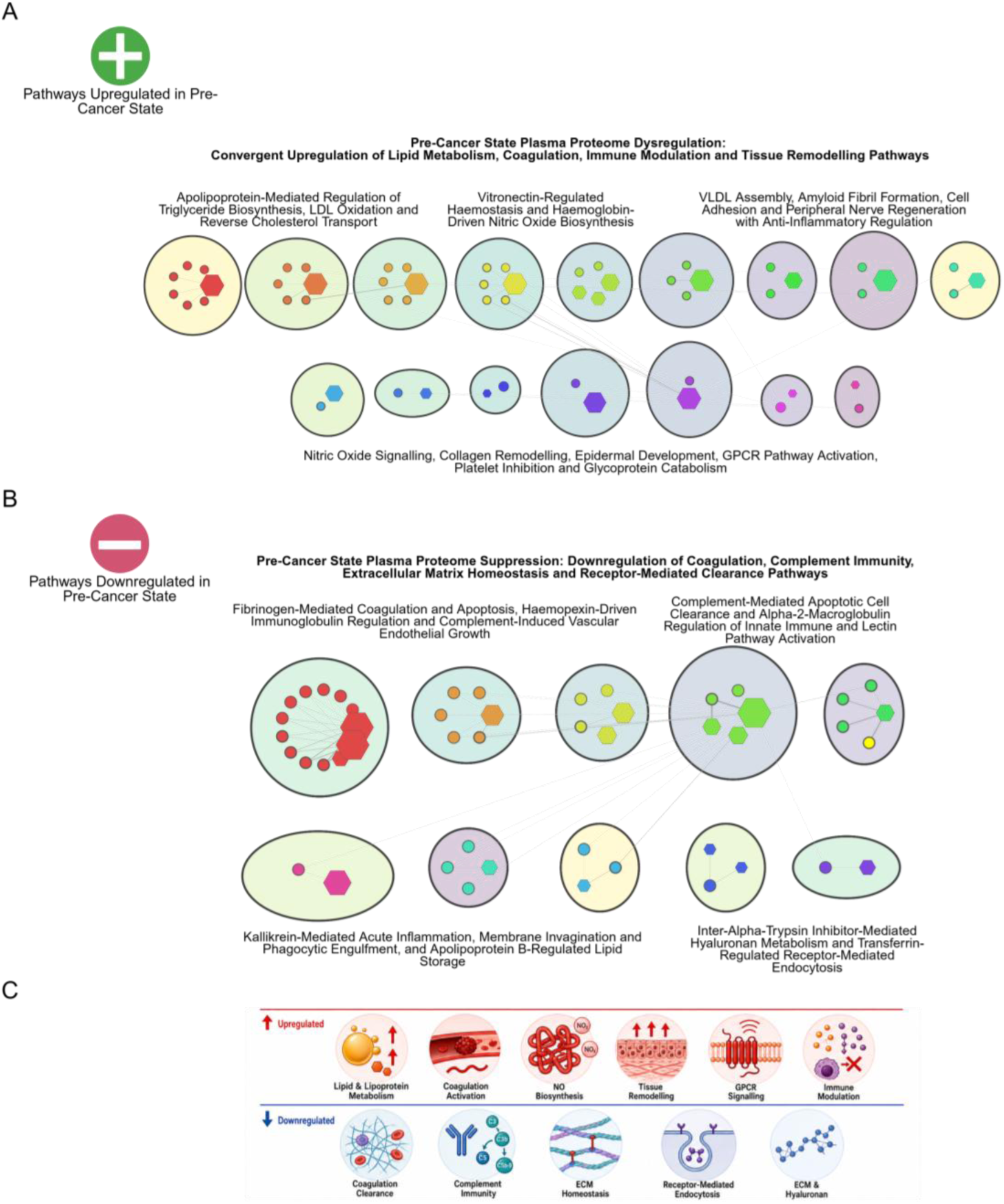
Systems-level pathway network architecture of the pre-cancer plasma proteome. a, GO Biological Process enrichment network of proteins upregulated in pre-cancer relative to healthy controls (“Pathways upregulated in pre-cancer state”), showing convergent enrichment for lipid and lipoprotein metabolism, coagulation activation, nitric oxide biosynthesis, tissue remodelling and GPCR signalling pathways. Nodes represent enriched GO terms; edges represent gene-set overlap; node size reflects gene count per term. b, GO Biological Process enrichment network of proteins downregulated in pre-cancer relative to healthy controls (“Pathways downregulated in pre-cancer state”), showing coordinated suppression of humoral immunity, complement activity, coagulation-associated defence, extracellular-matrix organisation and receptor-mediated clearance. c, Pictorial summary of the directional pathway changes in the pre-cancer plasma proteome. Upregulated pathways (red row): Lipid and Lipoprotein Metabolism, Coagulation Activation, NO Biosynthesis, Tissue Remodelling, GPCR Signalling and Immune Modulation. Downregulated pathways (blue row): Coagulation Clearance, Complement Immunity, ECM Homeostasis, Receptor-Mediated Endocytosis and ECM and Hyaluronan metabolism.

**Supplementary Fig. 8.**
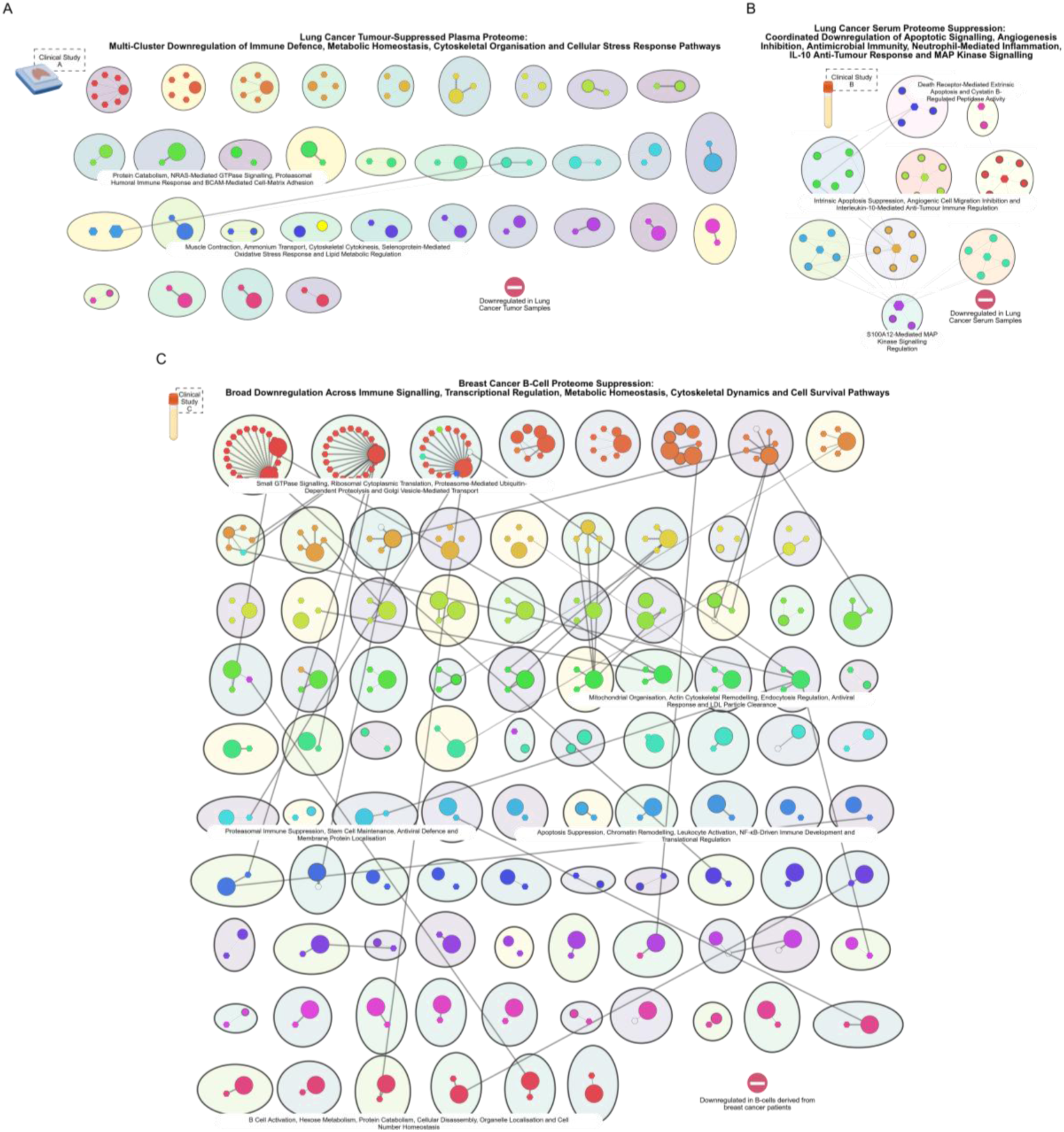
External lung and breast cancer proteomics datasets corroborate suppression of immune defence, cytoskeletal integrity and cellular stress response pathways in established cancer, extending the biological relevance of the exercise-derived signatures. a, GO Biological Process enrichment network downregulated in lung cancer tumour tissue versus healthy lung tissue (dataset PXD066683; combined_protein.tsv), showing multi-cluster downregulation of immune defence, metabolic homeostasis, cytoskeletal organisation and cellular stress response pathways. b, GO Biological Process enrichment network downregulated in lung cancer serum samples versus controls (dataset PXD034221), showing coordinated downregulation of apoptotic signalling, angiogenesis inhibition, antimicrobial immunity, neutrophil-mediated inflammation, IL-10 anti-tumour response and MAP kinase signalling. c, GO Biological Process enrichment network downregulated in B-cells derived from breast cancer patients (dataset PXD052185), showing broad downregulation across immune signalling, transcriptional regulation, metabolic homeostasis, cytoskeletal dynamics and cell survival pathways.

## Online methods

### Study overview and cohort framework

This study used serum samples from the Singapore Longitudinal Ageing Studies (SLAS I and II; n=6,050), a population-based cohort of community-dwelling adults in Singapore aged >=55 years at baseline, with longitudinal follow-up and standardized clinical assessments; SLAS is registered at ClinicalTrials.gov (NCT03405675). Prediagnostic pre-cancer samples were defined as sera collected from participants who were cancer-free at blood draw and subsequently diagnosed with cancer during follow-up. The analytical dataset and subgroup composition are summarized in Table 1. In this manuscript, structured exercise refers to supervised, protocol-prescribed training interventions (Malnutrition and Frailty Intervention (MFI) and Breast Cancer Exercise Intervention (BREXINT)), whereas unstructured vigorous physical activity refers to unsupervised, self-reported free-living activity behavior in the Frequent Exerciser cohort. Thus, the structured arm represents intervention-based exercise exposure, while the unstructured arm represents habitual community activity exposure outside a prescribed training protocol. Where applicable, the low-activity comparator is labeled no exercise for brevity; this denotes lower vigorous activity exposure rather than complete inactivity.

**Table 1.**
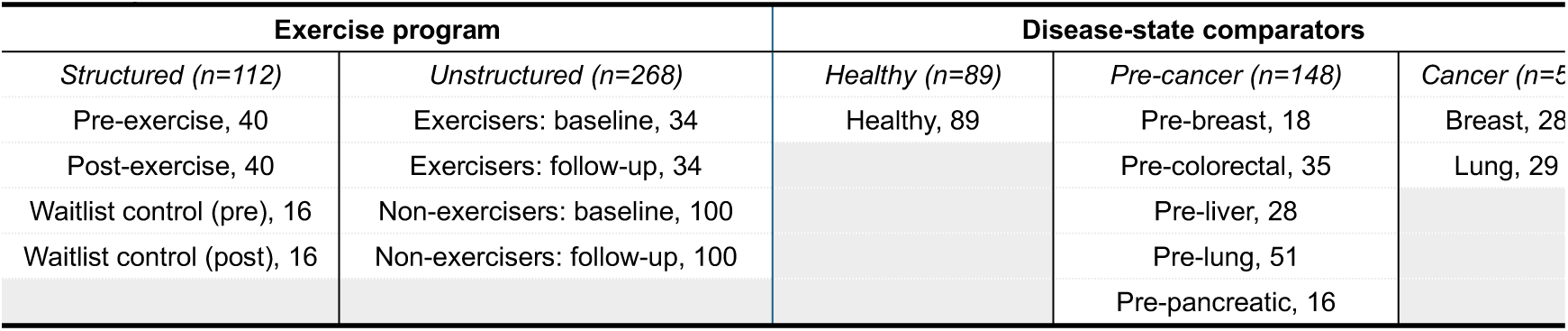
Cohort Composition.

### Structured exercise program cohorts

The structured exercise arm combined two Singapore intervention cohorts: MFI and BREXINT of ClinicalTrials.gov: NCT05957068. BREXINT was a 24-week randomized exercise trial in recovered breast cancer patients; all participants had undergone curative breast surgery and, where indicated, had completed chemotherapy with or without radiotherapy before study entry. For the present proteomic analysis, the BREXINT subset included three participants with paired sera (6 serum samples). Detailed cohort and protocol methods are provided in the referenced reports (MFI; BREXINT). The MFI program is contextualized within Singapore’s broader frailty-intervention clinical trial landscape, including NCT05007353, NCT06753643, and NCT07525583.

### Cancer patient cohort

The cancer patient cohort included patients with breast and lung cancer recruited at Singapore General Hospital between January 2005 and December 2015 and was used as contextual malignant-state biology. Cancer status, clinical variables, demographic variables, and tumor type were abstracted from medical records.

### Unstructured exercise program cohort (Frequent Exerciser cohort)

In the unstructured cohort, exposure was defined as self-reported, unsupervised free-living physical activity (that is, activity outside a supervised intervention), assessed at baseline and follow-up. Within this framework, exercise is considered a structured subset of physical activity, whereas the unstructured cohort captures habitual free-living behavior. Vigorous activity was operationalized as average daily duration (hours/day) on weekdays and weekend days, and participants were stratified using wave-specific 75th-percentile thresholds into longitudinal patterns (persistent low, baseline-only high, follow-up-only high, persistent high). To contextualize exposure, lifestyle behavior was recorded using standardized nurse-administered questionnaire items, including regular exercise (i.e., 2-3 times per week), fitness activities (calisthenics/aerobic/jogging/bicycle riding), walking, active sports (e.g., swimming, tennis, badminton, bowling, golf), and Taiji (body-mind exercise), each coded as Never (<1/month), Sometimes (>=1/month to <1/week), or Often (>=1/week). In figures and main text, the low-activity comparator is labeled no exercise for brevity; this label indicates lower activity exposure rather than complete inactivity. Accordingly, habitual denotes repeated exposure patterns across baseline and follow-up, whereas vigorous exposure is defined by self-reported duration rather than a single frequency threshold.

### Clinical phenotyping and derived indices

Clinical chemistry, hematology, and anthropometry were obtained at baseline and harmonized before analysis, including total cholesterol, LDL cholesterol, HDL cholesterol, triglycerides, fasting blood glucose, albumin, creatinine, estimated glomerular filtration rate, hemoglobin, hematocrit, red-cell indices, white blood cell count, differential leukocyte counts (neutrophils, lymphocytes, monocytes, eosinophils, basophils), platelet count, mean platelet volume, age, height, weight, body mass index (BMI), waist circumference, and hip circumference. Derived indices were then recalculated directly from these base variables using fixed formulas. Immune-inflammatory indices were defined as neutrophil-to-lymphocyte ratio (NLR = Neutrophils count / Lymphocytes count), platelet-to-lymphocyte ratio (PLR = Platelet count / Lymphocytes count), lymphocyte-to-monocyte ratio (LMR = Lymphocytes count / Monocytes count), and systemic immune-inflammation index (SII = Platelet count × Neutrophils count / Lymphocytes count). Cardiometabolic indices were defined as LDL:HDL ratio (LDL / HDL), TC:HDL ratio (TC / HDL), non-HDL cholesterol (TC - HDL), TG:HDL ratio (TG / HDL), atherogenic index of plasma (log10(TG / HDL)), Castelli Index I (TC / HDL), Castelli Index II (LDL / HDL), atherogenic coefficient ((TC - HDL) / HDL), and glucose:HDL ratio (Fasting Blood Glucose / HDL). Additional composite indices included creatinine:BMI ratio (Creatinine / BMI), RDW:hemoglobin ratio (Red Cell Distribution Width / Hemoglobin), and RDW:MCV ratio (Red Cell Distribution Width / Mean Corpuscular Volume). All derived features were appended to the analysis matrix and integrated with molecular and lifestyle variables for downstream modeling and visualization.

### Olink proteomics study cohort

The Olink Explore 3072 platform was used to profile plasma proteomes from 53,014 UK Biobank participants, measuring 2,923 analytes (mapping to 2,941 protein targets), with detailed assay and processing procedures described elsewhere. Cancer status was defined using ICD-10 hospital records (Field 41202), focusing on colorectal (C18-C21), lung (C34), breast (C50), pancreatic (C25), and liver (C22) cancers; participants without any recorded cancer diagnosis were classified as healthy controls. Physical activity variables were extracted from touchscreen questionnaire fields and grouped into unstructured and structured domains, where unstructured activity included walking (Field 971), light DIY (Field 1011), and heavy DIY (Field 224), and structured activity included strenuous sports (Field 991) and other exercise (Field 3637). To maximize exposure contrast, only extreme frequency categories were retained, with “Every day” defined as active and “Once in the last 4 weeks” defined as inactive, while intermediate frequencies were excluded to reduce exposure misclassification. After phenotype filtering and integration with the proteomic matrix, the final analytic dataset comprised 9,824 samples, which were used for external direction-of-change comparison with prioritized proteins.

### Serum processing and proteomics

For protein extraction, 50 ul serum was mixed with 1,700 ul of 75% acetonitrile (non-acidified) following the referenced precipitation workflow, then processed using the high-abundance protein depletion strategy. Peptides were analyzed by nanoLC-MS/MS using a Vanquish Neo system coupled to an Orbitrap Eclipse Tribrid mass spectrometer (Thermo Fisher Scientific) operated in positive-ion NSI mode. LC separation used direct injection in nano-capillary flow regime with a 75 µm inner-diameter, 50 cm analytical column; autosampler temperature was 7 °C; mobile phases were A (H_2_O) and B (80% Acetonitrile). The run time was 75 min at 0.300 µl/min with gradient 0-60 min to 40% B, 68 min to 50% B, 70 min to 100% B, and hold to 75 min (0.400 µl/min at run end), followed by post-run equilibration; loading/equilibration were pressure-controlled at 1200 bar. Additional autosampler settings included draw speed 0.200 µl/s, draw delay 2.0 s, dispense speed 5.000 µl/s, weak wash 5.0 s at 80.0 µl/s, strong wash 3.0 s at 80.0 µl/s, and wash mode after draw. Acquisition was DDA (cycle time 3 s, 0-75 min) with MS1 Orbitrap resolution 60K, m/z 350-1550, AGC target 400,000, max IT 100 ms, RF lens 55%, profile data; precursor filters included MIPS peptide mode, charge states 2-7 (undetermined excluded), and minimum intensity threshold 25,000; dynamic exclusion was set to exclude after 1 occurrence for 60 s with +/-10 ppm tolerance, isotope exclusion, exclude-within-cycle, and single-charge-state dependent scanning per precursor; MS2 used quadrupole isolation window 1.2 m/z, HCD stepped NCE 28/30/32, Orbitrap resolution 50K, AGC target 75,000, max IT 100 ms, first mass 110, one microscan, profile mode.

### Protein identification, quantification, and matrix generation

Raw files were processed in FragPipe v23.1 (MSFragger v4.3, MSBooster v1.3.17, Percolator v3.7.1, Philosopher v5.1.2, IonQuant v1.11.11). Database searching used a combined UniProt-OpenProt human FASTA with decoy/contaminant entries (decoy prefix rev_), semi-tryptic trypsin specificity (KR; <=2 missed cleavages), 20 ppm precursor and fragment tolerances, isotope error 0/1/2, precursor mass window −20 to +20 ppm, and enabled mass calibration (calibrate mass=2). Carbamidomethyl cysteine (+57.02146) was set as a fixed modification, with methionine oxidation (+15.9949; <=3 per peptide) and protein N-terminal acetylation (+42.0106; <=1 per peptide) as variable modifications. PSMs were rescored with Percolator and assembled in Philosopher at 1% FDR (picked/grouped) after contaminant removal. IonQuant LFQ was performed using match-between-runs (10 ppm m/z, 0.4 min retention time, 0.05 ion mobility), LFQ/maxLFQ, requantification, and ion-level FDR 0.01. Sample tracking and annotation were managed through the FragPipe manifest (cohort/group/replicate metadata), harmonized using experiment_annotation.tsv and SDRF files, with downstream provenance recorded in file list proteinprophet.txt and file list ionquant.txt. MSBooster incorporated retention-time and spectral prediction via Koina (best-model search enabled), and rescored identifications were propagated to IonQuant and MSstats-compatible outputs. Protein abundance matrices were then log2-transformed, median-normalized, and filtered to remove decoy/contaminant entries before generation of the curated analysis matrix.

### Integrated analytical workflow, reproducibility, and data/code availability

Exercise-hit proteins were identified separately for structured and unstructured programs using the paired within-subject workflow implemented in SLAS Final Main Script.R. Layout rows were mapped to matrix sample columns using a dual-overlap strategy (direct Protein Matrix Mapping ID mapping versus reconstructed Group row index mapping), and participant IDs were harmonized to a stable participant key (Clinical Base) by removing trailing A/B suffixes. True pre/post pairs were then generated within each arm by inner joining on Clinical Base. Protein-level testing was restricted to mapped samples, with optional missingness filtering (default: retain proteins with <=20% missing values), and statistical testing required at least three complete pairs per contrast. For each protein, mean pre, mean post, delta (post - pre), paired p-value, and direction were computed in intervention and comparator arms using paired two-sided t-tests (alpha = 0.05). HIT assignment followed two prespecified modes: Mode A (intervention significant and not reproduced as a same-direction significant comparator effect) and Mode B (intervention stable by p-value and optional delta-stability gate, default |delta| <= 0.20, with significant comparator change). With both modes enabled, final HITs were defined as proteins meeting either mode; structured-only, unstructured-only, and overlap HIT sets (set intersection) were exported for downstream analyses. Exercise-derived signatures were then projected into healthy, pooled pre-cancer, and cancer comparator spaces to assess separation, gradient structure, and longitudinal movement. The predefined workflow included 3D PCA, row-wise z-scored heatmaps, top-hit boxplots, composite signature scoring, volcano/delta summaries, and ROC/AUC modeling. For ROC analyses, predictors were zero-imputed and row-z-scored, top proteins were ranked by univariate AUC gain, and repeated stratified cross-validation was used for combined and single-protein models (including a combined model with selected clinical covariates where specified in-script). Differential comparisons in summary outputs used Welch-type two-group contrasts and ANOVA/Tukey-style procedures where applicable within the scripted pipeline.

Pathway and network interpretation used GO Biological Process enrichment (clusterProfiler) on program-level HIT sets, with Cytoscape/py4cytoscape automation to generate protein-pathway bipartite networks. Inputs were filtered by adjusted significance (default adjust < 0.05) and minimal topology constraints before expansion to protein-pathway edges; edge magnitude was encoded as −log10(pvalue). Node attributes included node type, degree, and enrichment metadata (including fold-enrichment where available), and visualization rules scaled node/edge aesthetics by topology and significance. To evaluate external robustness at the direction-of-change level, cross-platform concordance analysis was performed between Olink and LC-MS/MS on UniProt-matched proteins (isoform suffix removed), with non-finite values imputed to zero before platform-wise z-scoring. Olink deltas were computed as mean post - pre within strata; LC-MS/MS deltas were computed as mean paired post - pre within exercise arms. Scatter plots displayed all mapped proteins with finite deltas in high strata, colored by same-versus opposite-direction change; annotation statistics were interpretive and not used as plotting inclusion gates. Concordant-protein boxplots were generated from proteins with non-zero, same-direction deltas on both platforms, showing four-arm z-score distributions (Olink pre/post; LC-MS/MS pre/post paired) with overlaid mean trend segments. Reproducibility was enforced through version-controlled runtime checks and structured output manifests. Analyses and figure generation were executed through SLAS_Final_Main_Script.R and companion scripts, with pinned package/runtime metadata, section-level logs, QC/sanity tables, generated-file manifests, and source-data exports written into numbered output directories. Exported artifacts included HIT tables/lists, mapping and pairing tables, projection matrices, ROC inputs/curves/summaries, pathway/network tables, concordance outputs, figure source tables, and reproducibility documentation for end-to-end traceability. All source data underlying main and supplementary figures are provided in Source Data files; cohort-level sharing is governed by institutional approvals and consent constraints. Code for data processing, statistical analysis, and figure generation is available from the corresponding author on reasonable request and/or via the designated project repository.

### Figure preparation

Conceptual schematic illustrations and pictorial graphical elements were generated with the assistance of a generative artificial intelligence image tool (ChatGPT, GPT-5; OpenAI) and were subsequently reviewed, edited and assembled by the authors. No experimental data, quantitative plots, statistical analyses or data-derived figures were generated or altered using generative AI; all such figures were produced directly from the underlying data using the analysis code described above.

## Acknowledgements

We thank the iCORE Proteomics platform (formerly the Advanced Proteomics Platform, A∼PROMPT), Agency for Science, Technology and Research (A*STAR), Singapore, for mass spectrometry and proteomics services and technical support. This work was supported by the Singapore National Medical Research Council (OFLCG23may-0039, SYMPHONY 2.0; OFLCG24MAY-0028, CLARION; OFLCG24may-0025, Gastric LCG; CIAINV25jan-0001, DATANG; MOH-001067, Liver LCG PLANET 2.0; MOH-001610-00, Iron/CRC IRG), the A*STAR GAP Funding (I24D1AG059, EDDC EBC-129) and the Industry Alignment Fund– Industry Collaboration Fund (I2001E0065, Lucence).

## Notes

### Competing Interest Statement

The authors have declared no competing interest.

### Author Declarations

The Institutional Review Board of the National University of Singapore gave ethical approval for this work (Singapore Longitudinal Ageing Studies; reference 04-140). All participants provided written informed consent.

